# No Significant Difference in Viral Load Between Vaccinated and Unvaccinated, Asymptomatic and Symptomatic Groups When Infected with SARS-CoV-2 Delta Variant

**DOI:** 10.1101/2021.09.28.21264262

**Authors:** Charlotte B. Acharya, John Schrom, Anthea M. Mitchell, David A. Coil, Carina Marquez, Susana Rojas, Chung Yu Wang, Jamin Liu, Genay Pilarowski, Leslie Solis, Elizabeth Georgian, Maya Petersen, Joseph DeRisi, Richard Michelmore, Diane Havlir

**Affiliations:** Genome Center, University of California, Davis, USA; Unidos en Salud, San Francisco, CA, USA; Chan Zuckerberg Biohub, and Dept of Biochemistry & Biophysics, University of California, San Francisco, USA; Department of Medicine, Division of HIV, Infectious Disease and Global Medicine, University of California, San Francisco, USA; Joint UCB/UCSF Bioengineering Program, University of California, Berkeley and University of California, San Francisco, USA; The Public Health Company, Oakland, California, USA; School of Public Health, University of California, Berkeley, USA

**Keywords:** asymptomatic testing, COVID-19, Ct-value, SARS-CoV-2, Delta variant

## Abstract

We found no significant difference in cycle threshold values between vaccinated and unvaccinated, asymptomatic and symptomatic groups infected with SARS-CoV-2 Delta. Given the substantial proportion of asymptomatic vaccine breakthrough cases with high viral levels, interventions, including masking and testing, should be considered for all in settings with elevated COVID-19 transmission.

## Background

Vaccines reduce infection, severe disease, and death from SARS-CoV-2 (COVID-19) [1], yet breakthrough cases occur [2]. Several reports show no difference in cycle threshold values (Ct-values) between vaccinated and unvaccinated individuals [2, 3, 4]; however, others have suggested that breakthrough infections, particularly among asymptomatic individuals, have a lower viral load and therefore may be less likely to result in transmission [5, 6].

Effective epidemic control requires contemporary data to guide public health mitigation measures. Here, we report on Ct-values among fully vaccinated and unvaccinated individuals, asymptomatic and symptomatic at time of testing, during a period of high transmission of the Delta variant in two distinct populations: a Unidos en Salud (UeS) community-based site in the Mission District of San Francisco and Healthy Yolo Together (HYT) asymptomatic testing through the University of California (UC), Davis.

## Materials and Methods

### Study Populations

Data was collected on individuals who voluntarily sought testing for SARS-CoV-2 from two demographically distinct populations in California during a two-month period from June 17 to August 31, 2021, during which Delta was the predominant variant.

#### HYT

As part of the response to the COVID-19 pandemic, UC Davis deployed an extensive free asymptomatic testing program that included the City of Davis and Yolo County (<u>Healthy Yolo Together</u>). Asymptomatic individuals over the age of 2 were eligible for testing. Asymptomatic cases were classified as individuals not reporting symptoms at the time of testing. Samples were collected through a supervised method in which individuals transferred their saliva into a barcoded tube (<u>COVID-19 Testing | Campus Ready</u>). Smaller numbers of symptomatic individuals were processed using a different workflow and an antigen test; therefore, they were not included in this study.

#### UeS

The study population included individuals who sought SARS-CoV-2 testing at the UeS walk-up site, an ongoing academic (UC San Francisco, CZ Biohub, and UC Berkeley), community organization (Latino Task Force), and government (SFDPH) partnership. The outdoor, free BinaxNOW™ testing site was located at a public transport and commercial hub in the Mission District, a setting of ongoing transmission in San Francisco [7]. Individuals one year of age and older, with or without symptoms, were eligible for testing.

### Measurements

Infections were classified as breakthrough infections if the individual was fully vaccinated (two weeks following receipt of all vaccine doses). Individuals that had had only one dose or were tested within two weeks of the second dose, in the case of Pfizer and Moderna vaccines, were not included in the analysis.

#### HYT

Demographic information was collected from individuals at the time of registration. Vaccination status information was obtained at the time of contact tracing and confirmed in the California Vaccine Registry. Only confirmed, fully vaccinated individuals were used in the analysis; discordant samples, self-reported as vaccinated but unconfirmed, were treated as status unknown. Saliva samples from asymptomatic individuals were tested for the presence of the N1 and N2 regions of the viral nucleocapsid (N) gene using primers and probes described in the CDC 2019-Novel Coronavirus (2019-nCoV) Real-Time RT-PCR Diagnostic Panel, using IntelliQube high-throughput quantitative PCR instruments (LGC Biosearch Technologies). Ct-values were calculated with FastFinder software (<u>UgenTec | FastFinder</u>).

Genotypes of all N1/N2 positive samples were determined using RT-PCR SNP analysis at 11 loci diagnostic for variants of concern (<u>SARS-CoV-2 Variant ValuPanel assays | LGC Biosearch Technologies</u>). A subset of samples (39%) were also sequenced using the Illumina MiSeq sequencing platform. Consensus genomes were generated with Viralrecon2 and variants called in Pangolin version 3.1.11 and PLEARN-v1.2.66. Sequencing confirmed the variants called by genotyping.

#### UeS

Individuals provided demographic data and information on symptoms immediately prior to testing using BinaxNOW™ kits. COVID-19 vaccine status, including date of final shot, was obtained through the California Vaccine Registry. Anterior-nasal swab samples (iClean, Chenyang Global) collected by certified lab assistants from BinaxNOW positive individuals were placed in DNA/RNA Shield (Zymo, Inc.) and processed for qRT-PCR, genome recovery, and variant/lineage determination as previously described [8, 9]. Ct-values for the detection of N and E genes [8] were determined via the single threshold Cq-determination mode using Bio-Rad CFX Maestro v4.1 (Bio-Rad Inc). SARS-CoV-2 genomes were sequenced using the Illumina NovaSeq platform. Consensus genomes were generated via the COVID module of the IDseq pipeline (https://idseq.net) as described [9].

### Analysis

Ct-values were plotted, stratified by site; fully vs. not vaccinated; and symptom status. Partially vaccinated samples and stratification by age and vaccine type are reported in supplementary materials. Ct-values between strata were compared using a two sided t-test.

### Ethics Statement

#### HYT

The Genome Center laboratory that conducted COVID-19 testing was CLIA approved as an extension to the Student Health Center’s laboratory. The UC Davis IRB Administration determined that the study met criteria for public health reporting and was exempt from IRB review and approval.

#### UeS

The UC San Francisco Committee on Human Research determined the study met criteria for public health surveillance. All participants provided informed consent for testing.

## Results

A total of 869 samples, 500 from HYT and 369 from UeS, were included in the analysis. All analyzed samples from HYT were asymptomatic at the time of collection and 75% of the positive samples were from unvaccinated individuals (N=375). Positive samples from UeS were from both symptomatic (N=237) and asymptomatic individuals (N=132). The frequency of vaccine breakthroughs among the UeS samples (171 fully vaccinated, 198 unvaccinated) was greater than among the HYT samples, reflecting the different types of populations sampled. The Delta variant was the predominant variant detected in both populations (Supplementary Table 1).

There were no statistically significant differences in mean Ct-values of vaccinated (UeS: 23.1; HYT: 25.5) vs. unvaccinated (UeS: 23.4; HYT: 25.4) samples. In both vaccinated and unvaccinated, there was great variation among individuals, with Ct-values of <15 to >30 in both UeS and HYT data (Fig. 1A, 1B). Similarly, no statistically significant differences were found in the mean Ct-values of asymptomatic (UeS: 24.3; HYT: 25.4) vs. symptomatic (UeS: 22.7) samples, overall or stratified by vaccine status (Fig. 1B). Similar Ct-values were also found among different age groups, between genders, and vaccine types (Supplemental Figure 1).

**Figure 1.**
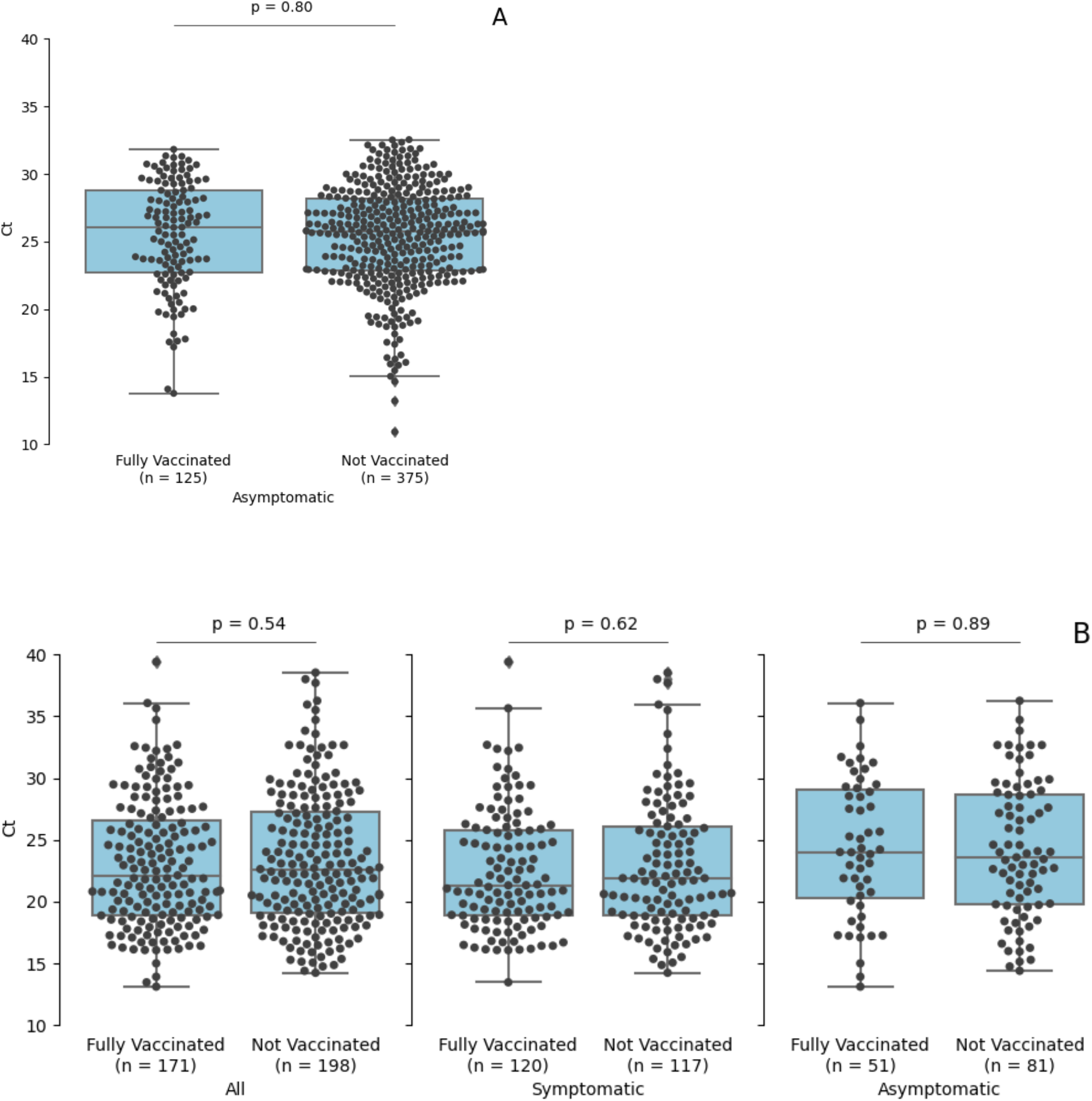
SARS-CoV-2 cycle threshold values in asymptomatic, symptomatic, vaccinated, and unvaccinated individuals in California. SARS-CoV-2 reverse transcription-polymerase chain reaction cycle threshold values for specimens from patients by vaccine status from Healthy Yolo Together (City of Davis and Yolo County, California) (Panel A) and from specimens by vaccine and symptom status from Unidos en Salud (Mission District, San Francisco, California) (Panel B). Box plots show first quartile, median, and third quartiles in shaded region; diamonds indicate outliers beyond 1.5 times the interquartile range; p-values were calculated with two-sided t-tests.

In all groups, there were individuals with low Ct-values indicative of high viral loads. A total of 69 fully vaccinated individuals had Ct-values <20. Of these, 24 were asymptomatic at the time of testing.

## Discussion

In our study, mean viral loads as measured by Ct-value were similar for large numbers of asymptomatic and symptomatic individuals infected with SARS-Cov-2 during the Delta surge, regardless of vaccine status, age, or gender. This contrasts with a large ongoing UK community cohort in which the median Ct-value was higher for vaccinated individuals (27.6) than for unvaccinated individuals (23.1) [5]. Also, a study from San Francisco reported that 10 fully vaccinated asymptomatic individuals had significantly lower viral loads than 28 symptomatic, vaccinated individuals [6]. Our study is consistent with other recent reports showing similar viral loads among vaccinated and unvaccinated individuals in settings with transmission of the Delta variant. In a Wisconsin study, Ct-values were similar and culture positivity was not different in a subset of analyses between 11 vaccinated and 24 unvaccinated cases [4]. In both Massachusetts and Singapore, individuals with vaccination breakthroughs caused by the Delta variant had similar Ct-values as unvaccinated individuals [3, 10]. Our findings are supported by consistency across large sample sets using different assays from two distinct locations.

A substantial proportion of asymptomatic, fully vaccinated individuals in our study had low Ct-values, indicative of high viral loads. Given that low Ct-values are indicative of high levels of virus, culture positivity, and increased transmission [11], our detection of low Ct-values in asymptomatic, fully vaccinated individuals is consistent with the potential for transmission from breakthrough infections prior to any emergence of symptoms. Interestingly, the viral loads decreased more rapidly in vaccinated than unvaccinated individuals in Singapore [3], suggesting that vaccinated individuals may remain infectious for shorter periods of time. Also, a retrospective observational cohort study of contacts of SARS-CoV-2-infected index cases in England documented reduced transmission from vaccinated individuals [12]. In our study, over 20% of positive, vaccinated individuals had low Ct-values (<20), a third of which were asymptomatic when tested. This highlights the need for additional studies of the immunological status of such vaccine escapes and how infectious they are. If such individuals carry high loads of active virus, asymptomatic vaccinated individuals may increasingly contribute to the ongoing pandemic as the proportion of vaccinated individuals grows.

Ct-values in some children under 12 who are not yet eligible for vaccination were also low. Twenty out of 109 (18.3%) children under 12 years of age had Ct-values <20, of which 14 were asymptomatic at the time of testing. Low Ct indicates that the children had high viral loads and were likely infectious. This emphasizes the value of regular, rapid testing for school children to detect infection early and block chains of transmission in settings where the Delta variant is circulating.

While vaccination remains the best protection against becoming infected and severe disease [12], the data gathered in this study during the surge of the Delta variant strongly support the notion that neither vaccine status nor the presence or absence of symptoms should influence the recommendation and implementation of good public health practices, including mask wearing, testing, social distancing, and other measures, designed to mitigate the spread of SARS-CoV-2.

## Data Availability

Data are available in the Supplementary Materials.

## Author Contribution Statement

JD, RWM, DH, and MP conceived the project. DC, CM, SR, DH, and GP helped collect the data. CA, AM, CYW, and JL helped perform the tests, genotyping, and sequencing. CA, JH, LS, JD, AM, CYW, JS, and JL prepared the data for publication. RM, EG, DH, MP, DC, JS, and JD contributed to the writing of the manuscript. All authors read and approved the final manuscript.

## Acknowledgements

Many people were responsible for collecting the samples, running the tests, performing the genotyping and sequencing, and processing the data as listed in Supplementary Table 2.

