## Supplementary Materials for "No Significant Difference in Viral Load Between Vaccinated and Unvaccinated, Asymptomatic and Symptomatic Groups When Infected with SARS-CoV-2 Delta Variant"

**Supplementary Table 1. Data summary of all samples collected and those included in this study.** The 869 samples collected between June 17 and August 31, 2021 from two populations in California and analyzed for this study. UeS: Unidos en Salud (UeS) community-based site in the Mission District of San Francisco, California. HYT: Healthy Yolo Together asymptomatic testing at the University of California, Davis, California.

| **Population** | **UeS** | **HYT** |
|---|---|---|
| Type of sample tested | Antigen test | Saliva |
| Patient status | Asymptomatic +Symptomatic | Asymptomatic |
| Total number of tests performed | 8,313 | 58,339 |
| Number of positive samples | 473 | 631 |
| Excluded due to missing symptom status | 7 | 0 |
| Excluded due to missing vaccination status | 97 | 21 |
| Excluded due to unknown/non-Delta lineage | -- | 110 |
| Number included in analysis | 369 | 500 |
| **Data Summary** | | |
| Percent positive (% of tested) | 5.69% | 1.08% |
| Percent of positives vaccinated (% of those included in analysis) | 46.3% | 25.0% |
| Percent symptomatic (% of those included in analysis) | 64.2% | 0% |
| Percent successfully genotyped or sequenced (% of positive samples) | 75.6% | 86.8% |
| Percent Delta (% of identified variants)* | 96.4% | 95.1% |

**Supplementary Table 2. Individuals involved in generating the data used this study.**

| Affiliation | Sector | Name |
|---|---|---|
| **Healthy Yolo Together** | Medical and Administrative Oversight | Ralph Green |
| | | Sheri Belafsky |
| | | Brad Pollock |
| | | Tod Stoltz |
| | | Ken Burtis |
| | Sample Collection | Karisa Gold |
| | | Sophia Haro |
| | Testing | Vanessa Rashbrook |
| | | Lutz Froenicke |
| | | Siranoosh Atari |
| | | Heather Matalka |
| | | Maria Kohler |
| | | Emily Kumimoto |
| | | Sherri Wykoff-Clary |
| | | Sheryl Bernauer |
| | | Nicole Slattengren |
| | | Kazuari Nozue |
| | | Helena Chang |
| | | Tommy Tran |
| | | Yin Lau |
| | | Melissa Horner |
| | Genotyping and Sequencing | Ana Menchaca |
| | | Monica Britton |
| | | Van Ly |
| | | Ryan Davis |
| | | Stephanie Liu |
| | Data Handling | Matt Settles |
| | | Adam Schaal |
| | | Jeff Trunnelle |
| **Unidos en Salud** | | |
| | Medical and Administrative Oversight | Salustiano Ribeiro |
| | | Douglas Black |
| | | Jen Nossokoff |
| | | Gabriel Chamie |
| | | Joselin Payan |
| | | Maria Powell |
| | | Shalom Bandi |
| | | Susy Rojas |

| | | |
|---|---|---|
| | | Rebecca Valencia |
| | Data Handling | James Peng |
| | | Simone Manganelli |
| | Community, Latino Task Force, and Unidos en Salud Staff | Valerie Tulier-Laiwa |
| | | Tracy Gallardo-Brown |
| | | Nelson Rivera |
| | | Alejandra Rodriguez |
| | | Chanagan Surrick |
| | | Jonathan Lemus |
| | | Margarita Sanchez |
| | | Cecilia Cuadra |
| | | Sonia Alvarenga |
| | | Patricia Lopez |
| | | Rosario Rodriguez |
| | | Glenda Cifuentes |
| | | Mario Delgado |
| | | Erica Vasquez |
| | | Xzalyn Hernandez |
| | | Patricia Murillo |
| | | Nayeli Veloz |
| | | Julie Satterfield |
| | | Helen Woldemariam |

A

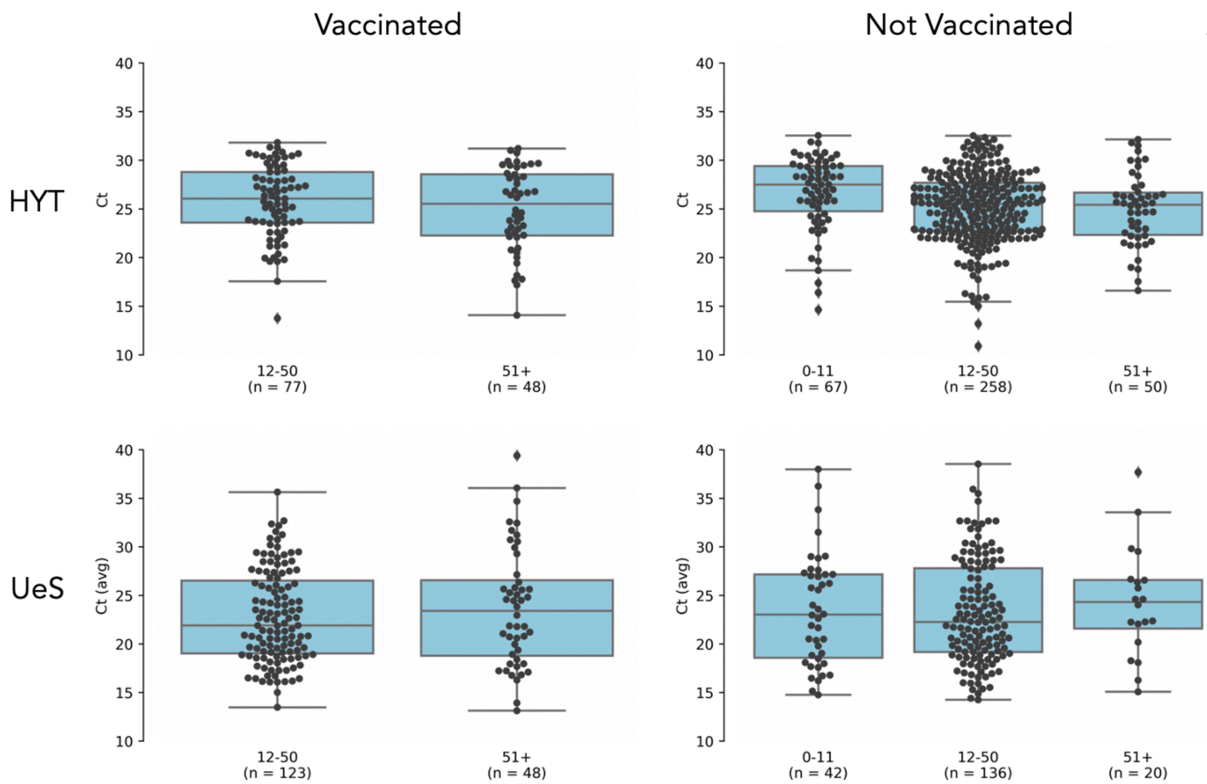
Vaccinated
Not Vaccinated
HYT
UeS
Ct
Ct (avg)
10
15
20
25
30
35
40
12-50
(n = 77)
51+
(n = 48)
0-11
(n = 67)
12-50
(n = 258)
51+
(n = 50)
12-50
(n = 123)
51+
(n = 48)
0-11
(n = 42)
12-50
(n = 136)
51+
(n = 20)

B

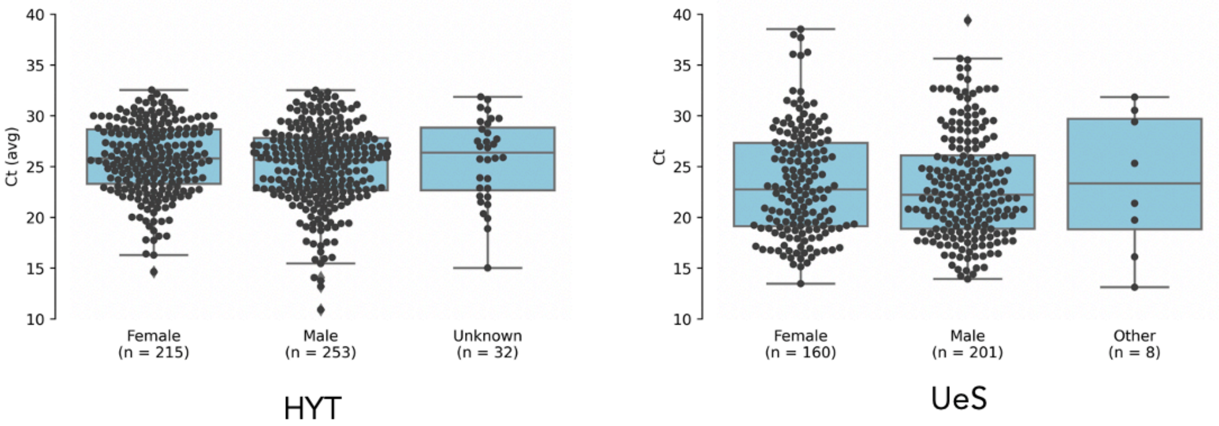
Ct (avg)
Ct
10
15
20
25
30
35
40
Female
(n = 215)
Male
(n = 253)
Unknown
(n = 32)
Female
(n = 160)
Male
(n = 201)
Other
(n = 8)
HYT
UeS

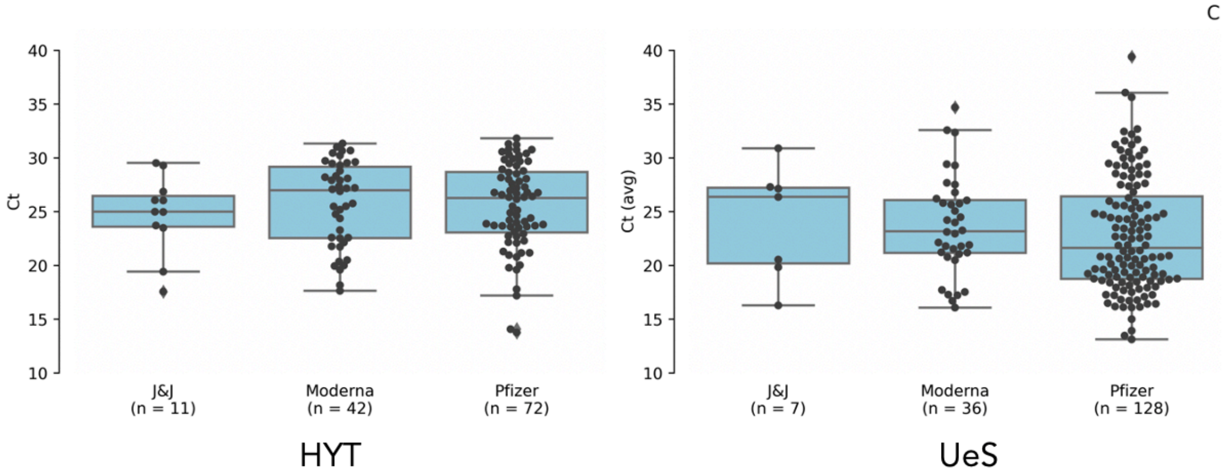

**Supplementary Figure 1. SARS-CoV-2 Ct-values stratified by age group, sex, and vaccine type.** SARS -CoV-2 reverse transcription-polymerase chain reaction cycle threshold values for specimens from patients by age and vaccine status from Healthy Yolo Together (City of Davis and Yolo County, California) and from specimens by vaccine and symptom status from Unidos en Salud (Mission District, San Francisco, California) (Panel A), by gender (Panel B), and by vaccine type (Panel C). Box plots show first quartile, median, and third quartiles in shaded region; diamonds indicate outliers beyond 1.5 times the interquartile range; p-values were calculated with two-sided t-tests.
